# Pharmacological correction of CFTR improves systemic inflammation and lung disease in Cystic Fibrosis but does not correct a basic defect in lung repair

**DOI:** 10.64898/2026.03.11.26348124

**Authors:** Nicola J Robinson, Gareth Hardisty, Jonathan Gillan, Robin Carajal Quisilema, Alfredo Montes Gomez, Debbie Millar, Stuart J Forbes, Robert D Gray

## Abstract

**Background:** Cystic Fibrosis (CF) is a lethal genetic condition affecting over 100,000 people worldwide, characterised by multi-organ dysfunction and a progressive lethal lung disease. The disease occurs due to faulty cystic fibrosis transmembrane conductance regulator (CFTR) ion channels effecting flow of chloride, bicarbonate and water out of cells. This causes thick mucus with repeated bacterial infections, systemic inflammation and a decrease in lung function.

CFTR modulator therapies have shown variable improvements in lung function and reduction in exacerbation frequency. Basal cells within the lung act as a stem cell for repair following injury and can repopulate the epithelial layer. This process is dysfunctional in CF causing progressive damage. Spontaneous lung repair is well described but not well characterised. Nothing is known about the effects of CFTR modulator therapy on these cells, but this could be of major consequence for people with CF (pwCF).

**Aims:** To determine the effects of CFTR modulator therapy on the activity of CF basal cells and relate this to progenitor function and to study the effects of CFTR modulators on systemic inflammation and clinical outcomes.

**Methods:** Clinical information, blood and nasal brushes were obtained from pwCF prior to commencing modulator therapy and at multiple time points up until 1 year of treatment. 10 pwCF were recruited to undertake thoracic CT scans pre-treatment and at 1 year of therapy. Nasal samples were used to isolate basal cells and serum to study systemic markers of inflammation. RNA sequencing of basal cells was undertaken by Ilumina Novoseq to a depth of 20 million read pairs and gene ontology analysis was performed. Functional assays of basal cell activity were carried out. Proteomic analysis and ELISAs were undertaken to determine changes in inflammatory cytokines within the serum across the first year of treatment. Quantitative results were generated by Lung Quantification (LungQ™) analysis with qualitative reports from independent radiologists. Results were compared with clinical outcomes.

**Results:** 110 pwCF were recruited in total who commenced a commercially available CFTR modulator therapy. Serum samples were collected from 77pwCF, nasal brushes obtained from 40 pwCF and 10 completed their CT scans following 1 year of highly effective CFTR modulator therapy.

Systemic IL-6, CRP and calprotectin (a biomarker of CF exacerbation) were all significantly reduced with highly effective CFTR modulator treatment.

Clinical results were in keeping with those seen in published CFTR modulator clinical trials with improvement in lung function, weight, and exacerbation frequency.

Subjective improvements were seen in all 10 CT scans following 1 year of modulator therapy. Significant reductions were seen in airway wall thickening and reduction in thoracic lymphadenopathy were also observed.

Basal cell RNA sequencing showed that the relative expression of 2570 genes were significantly different following treatment with CFTR modulators. Ontology analysis showed enrichment in multiple pathways including cilliagenesis and Notch signalling, a key pathway in lung tissue development and homeostasis. Functional assays exhibited a deficit in repair mechanisms of the CF basal cell compared to healthy controls, and reduction in progenitor function.

**Conclusions:** Although CFTR modulators improve multiple clinical and radiological outcomes, they also have impacts on basal cell function. There are however, limited impacts on systemic inflammation and more work is needed in this area to understand the disease process.

## Introduction

Cystic Fibrosis (CF) is a life limiting, multi-system, monogenic disease causing dysfunction of the chloride channel CFTR and subsequent mucous retention in the lung and chronic infection and inflammation leading to destructive lung disease^1,2^. Until recently, therapeutic intervention focussed on treating the symptomatic consequences of CF such as mucolytics to enhance mucous clearance and nebulised antibiotics to treat chronic infections leading to a high treatment burden that slowed but didn’t stop lung function loss^3–5^. CFTR dysfunction is also associated with a hyperinflammatory phenotype in immune cells, and people with higher levels of inflammation have a worse clinical outcome, suggesting a central role for inflammation in the progression of CF lung disease^6–10^. Over the past decade drugs targeting the basic defect in CF, abnormal CFTR protein, have been developed into life-changing treatments that partially reverse lung function loss in people with CF and are associated with improvements in overall health status^11–14^. It is unclear at present whether these improvements in lung function are due mainly to changes in mucous viscosity and clearance, or whether changes in inflammation have a positive impact on lung disease. Additionally, it is not clear whether the reversal of the basic defect in CF will lead to repair of already damaged lung or a persistent inflammatory lung disease. The introduction of these drugs into CF clinical populations offers a unique opportunity to study how correcting the basic defect in CF impacts lung disease, inflammation and tissue repair processes in a clinical population.

Work by our group and others have shown improvements in local and systemic inflammation and changes in inflammatory cell phenotype with a range of CFTR modulators^15–17^, with some studies also showing radiological improvements in lung disease^13,14,18^. The impact of correcting CFTR function on airway repair mechanisms is less clear, particularly when placed in the context of previous studies of the CF airway demonstrating increased numbers of basal cells (progenitor cells) but paradoxically less ciliated airways suggesting dysfunctional repair, which could be a consequence of fundamental defects in CF progenitor cells or the impact of inflammation as is seen in other diseases effecting the airway such as asthma and COPD.^19–23^ Taken together this suggests that correction of CFTR and reduction of inflammation could both be targets to repair lung damage in CF and secure long lasting benefits for lung health in CF.

Our present studies lever the sequential availability of 2 CFTR modulator drug treatments, first the dual combination Tezacaftor/Ivacaftor (TI) and next the triple combination Elexacaftor/Tezacaftor/Ivacaftor (ETI) to allow the study of inflammation in CF at different levels of CFTR correction. TI is a partial CFTR modulator, with a mean 4% improvement in percentage predicted forced expiratory volume in one second (ppFEV1) and decrease in sweat chloride of 10mmol/L^24^. ETI is more effective with a 14% mean improvement in ppFEV1 and reduction in sweat chloride of 40mmol/L^12^. We used the introduction of these treatments to undertake a series of observational experimental medicine studies to assess how CFTR correction effects inflammation (at different levels of CFTR augmentation), and the effects of CFTR correction on stem cell populations from the airway. In an additional cohort, we assessed radiological changes following CFTR correction and compared this directly with clinical parameters to phenotype deeply their response to ETI.

In summary our studies of clinical, radiological, inflammation and epithelial cell outcomes increases the understanding of how correcting CFTR may improve inflammation and lung health whilst highlighting areas of CF pathophysiology where further therapeutic development will be required

## Methods

Ethical approval was gained for enrolment of CF participants and healthy volunteers (IRAS IDs: 261543 and 286836) to obtain blood, nasal brushing samples and imaging.

### Flow Cytometry

300μl whole blood was separated into 3 tubes of 100μL. Antibodies were added (see Table 1 &2 online supplement) in 100μL staining buffer (2% FCS in PBS^-/-^) for 30 minutes at 4ᵒC. Following incubation, samples were lysed with 2mls FACSlyse (Biolegend) RBC lysis buffer for 10 minutes at room temperature and centrifuged at 300 x g for 5 minutes. Cells were washed in 2mls staining buffer and centrifuged again. This process was again repeated and cells were then fixed in 4% PFA (Sigma) in PBS^-/-^. Samples were run on a Fortessa LSR II (BD) within 2 days and analysed in FlowJo software (BD).

### Apoptosis assays

Neutrophils were harvested and centrifuged and washed then resuspended in binding buffer to 100µL. 5μL of Annexin V-FITC staining solution (Bio-techne). The mixture was incubated for 20 minutes at room temperature in the dark. A further 200μL of binding buffer was added to each tube and mixed. 1µL of Propidium Iodide was added just prior to analysis by flow cytometry on the NXT Attune.

### ELISAs

Serum samples from CF and healthy control samples were utilised. ELISAs were undertaken using Human IL-8, Human IL-6, Human IL-10 and Human TNF alpha Duoset ELISA kits (Biotechne) and calprotectin ELISAs were undertaken using Hycult calprotectin Elisa kits (Cambridge Bioscience). All samples were run in duplicates. All detection steps employed streptavidin-horseradish peroxidase (HRP) following attachment of biotinylated detection antibodies. TMB substrate (Invitrogen) was added per well and the plate was protected from the light for 5-30 minutes, depending on a colour change observed in the most highly diluted standard wells, at which point the reaction was stopped using KPL TMB stop solution (SeraCare) per well. Within 15 minutes, plates were read at 450nm and 570nm on The Synergy HT plate reader (Biotek). The 570nm background reading was subtracted from 450nm level and the mean of sample duplicates was taken and analysed using Graphpad Prism to interpolate the results to fit the standard curve.

### Cytokine bead array

Serum samples were run in duplicates. The LEGENDplex™ Human Inflammation Panel 1 (Biolegend) was used.

### Olink Proteomics

20 paired CF serum samples (before and after 1 year of ETI treatment) for proteomic analyses were analysed by OLINK proteomics using 2 panels (Olink Target 96 Inflammation and Olink Target 96 Organ Damage). Results were given by NPX value (log2). Bioinformatics by Olink utilised an adjusted p value of ≤0.05 as significant.

### CT scans

10 CFTR Modulator naïve CF participants (no previous generation CFTR modulator taken) with a single copy of the F508del mutation were recruited to undergo an inspiratory High-Resolution CT Thorax prior to commencing treatment and 1 year after commencing treatment with ETI. The pre and post treatment scans were analysed by 2 independent radiologists for qualitative analysis and then sent to Thirona for further Lung Quantification Analysis (LungQ™) using quantitative lung volume, airway and pulmonary arterial measurements to create a PRAGMA disease score for total disease, bronchiectasis and airways.

### Basal Cell Assays

**Basal cell counts**: At the time of seeding and first passage, cells were counted using a nucleocounter. Normalised rates of growth were calculated by number of cells at first passage, divided by number of cells at seeding, divided by time in days since seeding and displayed as million cells/day (Basal cell progeny per 100,000 cells at seeding from nasal brush).

**BRDU assay:** CF and healthy control basal cells were seeded at 30,000 cells/well in a pre-coated 24 well plate (Corning). Each sample was run in duplicate. 10µM BRDU labelling solution (Abcam) was added to the culture media and samples were grown for 24 hours. They were then fixed and stained with anti-BRDU antibody (Abcam) as and samples imaged. Cell numbers were counted, and results were expressed as a percentage of BRDU positive cells of all cells.

**Basal cell apoptosis rates:** 30,000 basal cells per sample well were seeded in 24 well plates and grown for 72 hours. Cells were harvested and centrifuged and resuspended in binding buffer to 100µL. 5μL of Annexin V-FITC staining solution (Bio-techne) was added to the tubes and mixed. The mixture was incubated for 20 minutes at room temperature in the dark. A further 200μL of binding buffer was added to each tube and mixed. 1µL of Propidium Iodide was added just prior to analysis by flow cytometry on the NXT Attune.

**Migration assay (wound closure):** Cells were grown to confluence and then a scratch was created down the centre of the well using a 200µL pipette tip. Samples were run in duplicate. Wells were imaged using the EVOS automated microscope at 20x magnification at 0 hours and 24 hours and stitched images were prepared with the results. Stitched images were analysed using Image J(FIJI) to calculate the area of the defect. Percentage reduction in defect area over 24 hours was quantified.

### Organoid formation and imaging

25,000 expanded basal cells were suspended in 50µL Matrigel on ice. This was pipetted into a dome on a pre-warmed 24 well non-adherent plate and incubated at 37°C for 30 minutes. For the first 7 days, 500µL of airway organoid seeding media (Stemcell Pnemacult Organoid media) was added to the well with media changes every 48-72 hours. This was then converted to Airway organoid differentiation media from day 7-28. Images were taken of the organoids at day 14 using the EVOS automated microscope at 10x magnification with stitched images to show the whole well. A sample of the 10x images were analysed with ImageJ (FIJI) to ascertain the 10 largest organoid areas within a 10x magnification frame and results expressed as a mean.

### RNA sequencing

9 modulator naïve CF participants were identified with samples prior to treatment and at 3 months of treatment. Their 3-month basal cells were grown for 5 days with ETI added to the media. 9 age and sex matched healthy controls were also identified and basal cells expanded.

1 million basal cells from each sample were mixed with 500 µL of Trizol (Invitrogen) and RNA was extracted using Quick RNA Miniprep Kit (Zymo Research). RNA quantity and quality was checked prior to sending to Genewiz for RNA sequencing. Library preparation and sequencing was carried out by Genewiz. Raw sequencing reads were assessed for quality using FastQC (v0.11.9) and summarised using MultiQC. Adapter sequences and low-quality bases were removed using Trim Galore (v0.6.7) with default parameters and post-trimming quality was re-assessed using FastQC and MultiQC. Trimmed reads were aligned to the GRCh38 human reference genome (GRC h38, patch 14)using STAR (v2.7.10a) with default settings. Gene-level quantification was performed using featureCounts (Subread v2.0.3) with Ensembl gene annotations, counting reads mapping uniquely to annotated exons and summarising at the gene level. Differential expression analysis was conducted in R (v4.3.0) using DESeq2 (v1.40.0). Raw count matrices were imported into DESeq2 and modelled using a negative binomial generalised linear model. Genes with low expression were filtered by requiring a minimum of 10 total counts across all samples. Differentially expressed genes were defined as those with Benjamini–Hochberg adjusted p-value < 0.05 and absolute log2 fold change > 0.5. Data exploration, visualisation, and differential expression analysis were performed using the Searchlight2 workflow in R with DESeq2-normalised expression values. Functional enrichment analysis of significantly upregulated and downregulated gene sets was performed using clusterProfiler (v4.8.0) and STRING (v11.5) via over-representation analysis, and terms with adjusted p < 0.05 were considered significant.

### Statistics

All statistics were analysed with Prism Graphpad. Statistical tests are documented for individual figures. Statistically significant results are denoted throughout this work as *=p<0.05 **=p<0.01, ***=p<0.001 and ****= p<0.0001. For correlation matrix of the deeply phenotyped patient sub-group; their individual parameters were collated and the difference in each parameter from 1 year of ETI treatment was compared with that of their pre-treatment values. The results were then compared using Prism Graphpad to analyse Pearson R values and significant p values for respective parameters.

## Results

110 people with CF (PWCF) were recruited over the study period. Due to Covid-19 restrictions, drug side effects, and engagement of PWCF at various points during the study period not all PWCF completed all study visits and sample collections. Demographics are presented in table 1. Assays performed and numbers of patients used for each assay are recorded in supplementary data table 3.

**Table 1:** Patient demographics.

|  |  |  |
| --- | --- | --- |
| **Total (N):** |  | **110** |
| **Sex:** | Male | 65 |
|  | Female | 45 |
| **Median Age :** |  | 29 |
| **Genotype:** | $\Delta F508/\Delta F508$ | 54 |
| | $\Delta F508/\text{other}$ | 51 |
|  | Other/Other | 5 |

### Partial CFTR correction with Tezacaftor/Ivacaftor (TI) combination had limited effects on systemic inflammation and immune cell phenotype

Despite modest changes in ppFEV1 with TI (2.5% mean improvement at 1 month and 2.9% at 3 months), there were very limited changes in markers of systemic inflammation. Despite examining an extensive panel of serum inflammatory cytokines with a multiplex immunoassay, only IL-6 demonstrated a statistically significant difference from healthy controls and decrease with TI treatment. Serum calprotectin, previously shown to be a major biomarker of CF exacerbation, didn’t change significantly with TI treatment at 3 months. Flow cytometry panel of neutrophil and monocyte maturity and activation markers was undertaken at baseline, 1 and 3 months of TI treatment (Figure S1). Significant increases were seen at baseline in CD66b, CD11b and CD35 in the CF cells compared to healthy controls but there was no significant difference in these following 3 months of TI treatment. CXCR2, a marker of neutrophil maturation, remained significantly reduced despite TI treatment. Taken together these results suggest that partial CFTR correction with TI is not sufficient to have a major impact on inflammation in pwCF.

**Figure 1A-C:**
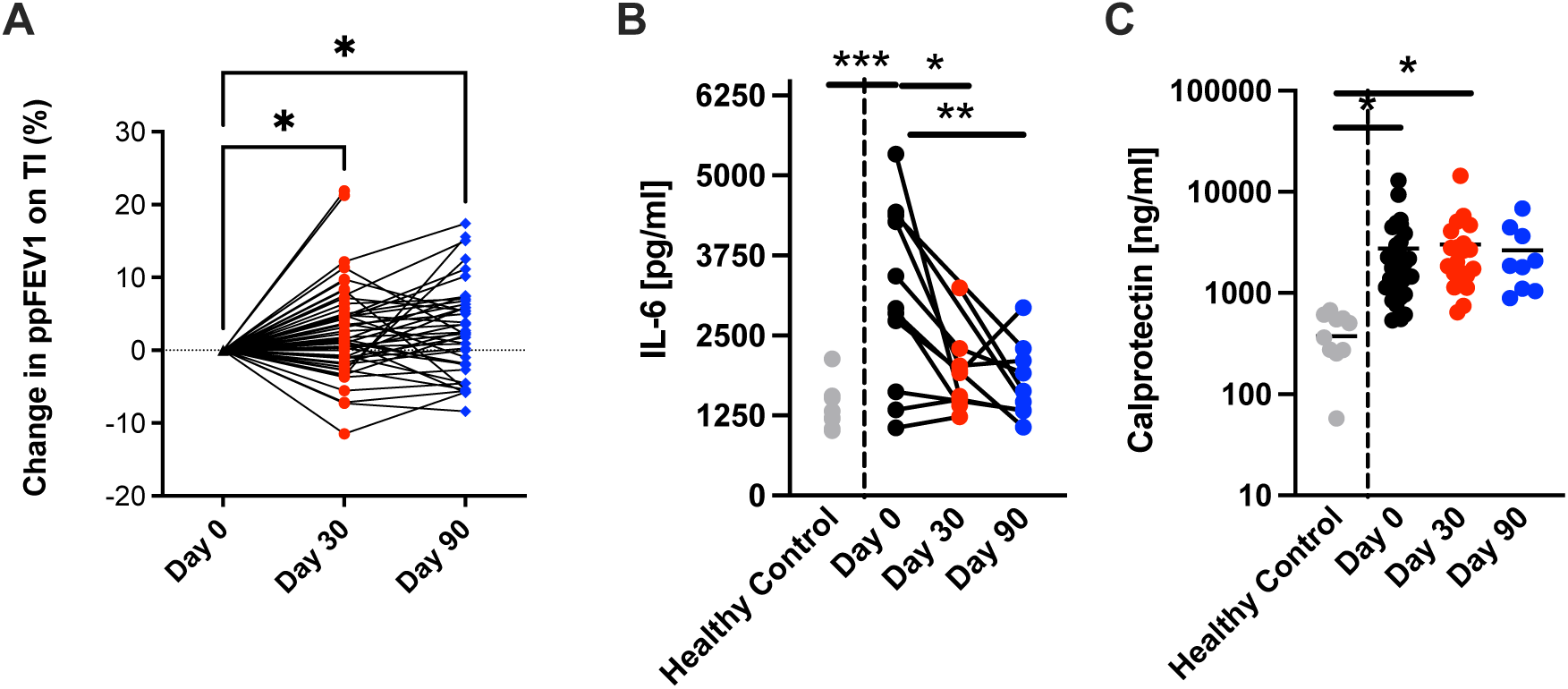
Systemic inflammation during TI therapy commencement. 1A demonstrates the change in ppFEV1 for individuals on TI over the first 3 months of treatment (n=51). 1B demonstrates the change in IL-6 measured by cytokine bead array comparing healthy serum to CF modulator naïve and following 30 and 90 days of TI therapy (n=9 healthy control and 12 CF paired samples pre and post treatment with TI). 1C shows the change in calprotectin, measured by ELISA comparing healthy serum to CF modulator naïve and following 30 and 90 days of TI therapy (n=11 healthy control, 31 pre treatment CF, 21 Day 30 TI CF and 9 Day 90 TI CF). Statistics with one way ANOVA with multiple comparisons where *≤0.05, **≤0.01, ***≤0.001, ****≤0.0001.

### Greater CFTR correction with ETI has significant effects on circulating immune cell numbers but modest effects on immune cell phenotype

Next, we assessed the effects of a more substantial correction of CFTR function in patients commencing ETI therapy. There was a significant reduction in global leukocyte counts after 3 months of ETI therapy. Prior to ETI treatment, 16.9% of PWCF had neutrophils and 29% monocytes above the upper limit of normal, reducing to 4.5% and 5.7% by the end of 3 months of ETI treatment. Males with CF (p=0.015) and those with moderate or severe lung disease (p=0.041) had a higher number of circulating monocytes. Neutrophilia was also more predominant in those with moderate or severe lung disease (p=0.005). Platelet counts also improved significantly over 3 months of ETI treatment with an increase of 10% of pwCF returning into the normal range.

Guided by previous data from TI treated PWCF we conducted multiparameter flow cytometry on neutrophils and additionally monocytes (having recently demonstrated that circulating monocytes in PWCF show higher levels of classical monocyte activation). Assays were conducted at 0 and 90 days of treatment demonstrated that circulating immune cells changed phenotype over the course of short-term treatment with ETI. Neutrophil activation and degranulation markers changed significantly with a reduction in CD35 and CD47 with increases in CD66b and CD88. An increase in CD16 staining in neutrophils suggesting increased maturity of circulating neutrophils following ETI.

Monocytes demonstrated a significant reduction in both CD11b and TNFR2 suggesting less activation, however a non-significant trend towards lower CD14 staining may suggest a shift from classically activated monocytes to a more intermediate phenotype.^9^

In summary our data from people taking ETI demonstrate a reduction in circulating myeloid cells associated with the innate immune response (neutrophils and monocytes) with neutrophils having a more mature phenotype and monocytes being less activated, suggesting a reduction in neutrophil release from bone marrow and a shift towards less classically activated monocytes in circulation.

### Specific serum markers of inflammation improve significantly on ETI in PWCF, even after only 1 month of treatment

Next, we measured a specific panel of inflammatory biomarkers based on previous studies of systemic inflammation in CF^8,16,25^ and data from our TI treated group. CRP, IL-6, IL-8, IL-10 and calprotectin were measured at one month and three months following treatment. There was a four-fold reduction in C-reactive protein in the initial 3 months after commencing treatment (p=0.0021). IL-6 was significantly reduced (p=0.0023) at 1 month and correlated with CRP reduction, however, there was no significant change seen in circulating levels of IL-8 or IL-10. Calprotectin, a biomarker of exacerbation risk, was significantly reduced (p=0.0011) after 3 months of ETI in patients. pwCF who were previously treated with TI and switched to ETI at the start of the study had lower levels of calprotectin at baseline, suggesting an impact of longer-term therapy with TI on calprotectin levels even before switching to a more effective CFTR therapy in contrast to the lack of effect with TI in the shorter term.

**Figure 2A-D:**
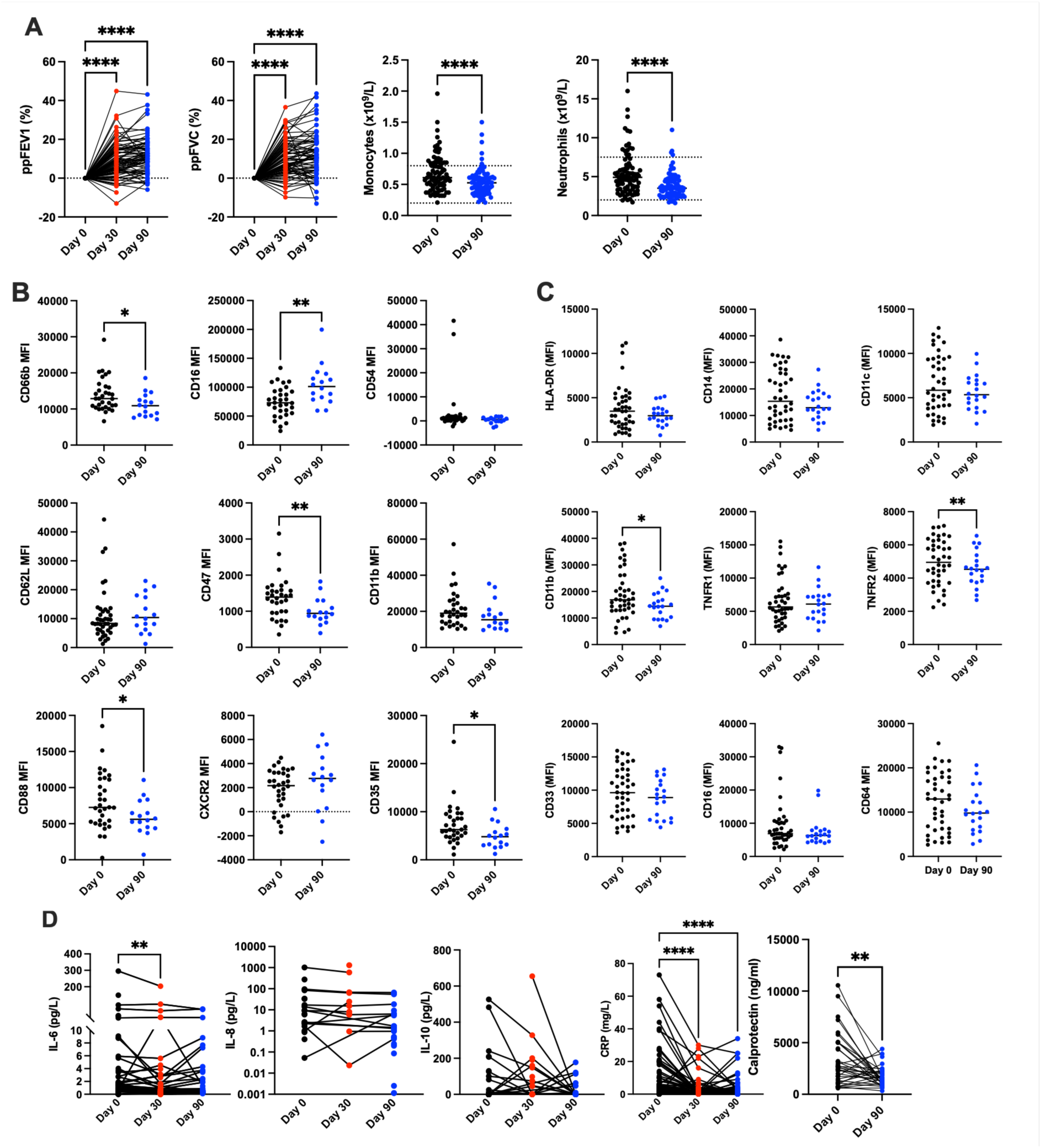
Despite significant improvements in lung function, ETI causes limited changes in systemic markers of inflammation over 3 months. 2A demonstrates the significant improvements in ppFEV1 and ppFVC (n= 89) and significant reduction in circulating neutrophils, monocytes, across the first 3 months of ETI therapy (n=89, dotted lines show the upper and lower limits of normal in the healthy population). 2B shows the mean fluorescence intensity (MFI) for multiple flow cytometry markers on neutrophils across the first 3 months of ETI therapy and with comparison to healthy controls (n=32 CF day 0 and 16 CF day 90). 2C shows the Mean Fluorescence intensity (MFI) for multiple flow cytometry markers on monocytes across the first 3 months of ETI therapy and with comparison to healthy controls (n=32 CF day 0 and 16 CF day 90). 2D shows the significant reduction in IL6 at 1 month (n=64 pwCF), and a significant reduction in CRP(n=75pwCF). IL-8 and IL-10 are unchanged in this cohort (n=64). Serum calprotectin is significantly elevated in pwCF and significantly reduces in this cohort with ETI treatment (n=34 pwCF). Statistics with Welch’s and paired t tests and one way ANOVA with multiple comparisons where *≤0.05, **≤0.01, ***≤0.001, ****≤0.0001.

### Longer-term ETI treatment (1 year of therapy) improves specific inflammatory markers in CF

By 1 year of ETI treatment, there was a significant improvement in lung function with a mean increase in ppFEV1 of 9.2% and a mean increase in ppFVC of 9.1%. We also saw a mean weight increase of 4.3kg over the first year of ETI treatment. Additionally, our cohort had a reduction in mean exacerbation frequency from 2.2 exacerbations/year to 0.6 exacerbations/year.

Given the sustained improvement in lung function, improved nutritional status and marked reduction in exacerbation, we assessed the longer-term effects of CFTR correction with ETI on systemic inflammation. Leucocyte counts were similar to those seen at 3 months but with an additional significant reduction in lymphocytes at 1 year.

Initially we utilised a 184-protein inflammation specific array (Olink) in a test set of 10 PWCF who were completely modulator naïve at baseline and after a year of ETI therapy. Remarkably only 3 proteins changed significantly after one year of treatment; namely IL-6, MMP-10 and IL-20 (although the array did not include key CF biomarker calprotectin). We then measured IL-6 and MMP-10 in our larger cohort of patients by ELISA and confirmed a reduction in these biomarkers at 1 year of therapy, additionally calprotectin also showed a sustained reduction at 1 year.

**Figure 3A-D:**
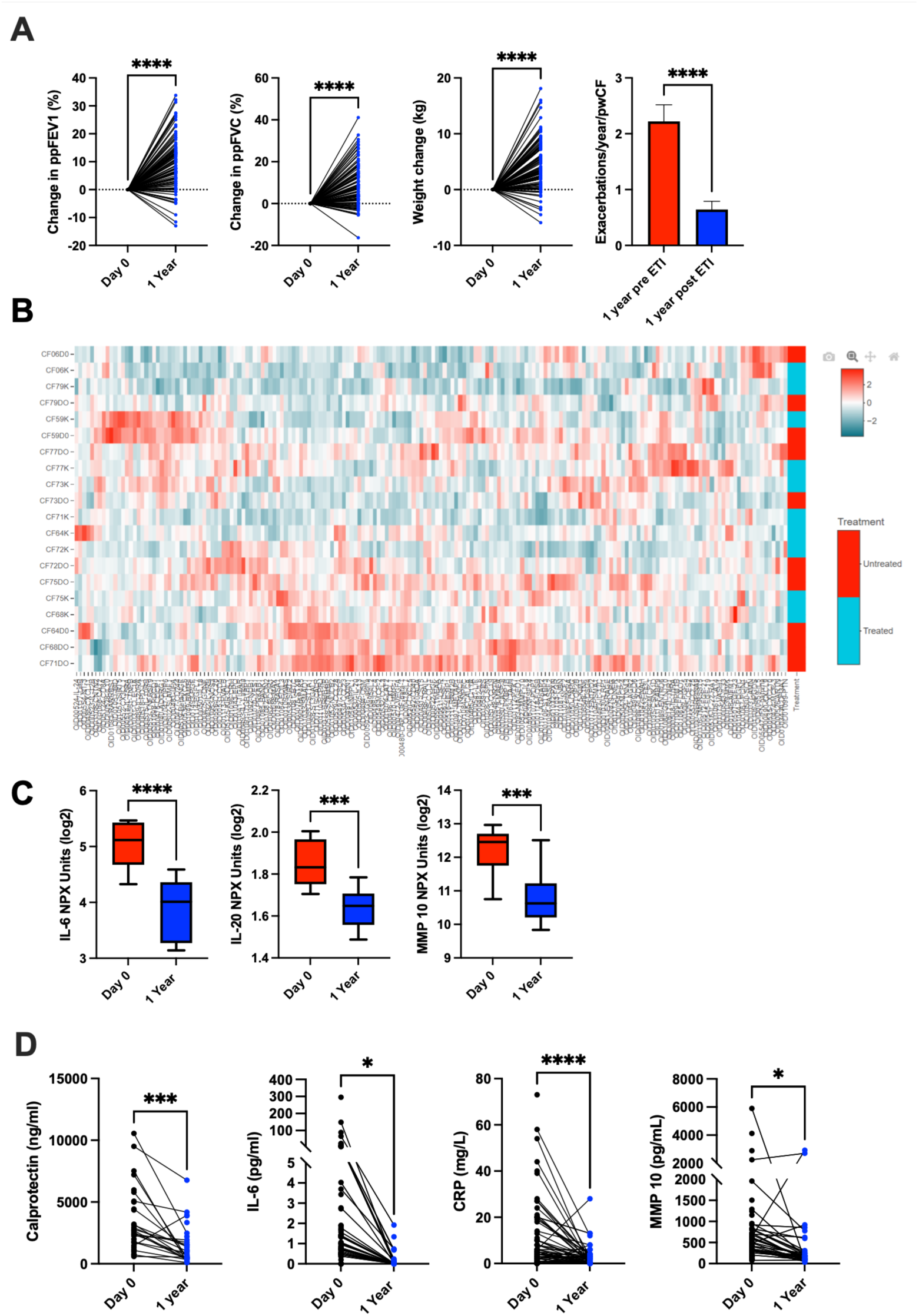
Systemic leucocyte and inflammatory responses to ETI are more marked in the longer term but no correlation is seen with an improvement in lung function or exacerbation frequency. 3A shows the clinical changes within the large cohort at 1 year, demonstrating the changes in ppFEV1, ppFVC, weight and exacerbation frequency (n=91) 3B shows a heatmap of 184 proteins assayed from CF serum before and after 1 year of ETI therapy within minimal changes in inflammation (n=10 pre and post ETI). 3C demonstrates the 3 significant results found from the proteomic analysis, IL-6, IL-20 and MMP-10 (n=10 pre and post ETI). 3D shows ELISA results across 1 year of treatment with ETI therapy showing significant reductions in Calprotectin (n=40), IL-6 (n=60 pre ETI and 40 1 year ETI), CRP (n=77) and MMP-10(n=60 pre ETI and 40 1 year ETI). Statistics with paired t tests for where *≤0.05, **≤0.01, ***≤0.001, ****≤0.0001.

### Long term ETI treatment is associated with changes in structural lung disease and reduction in multiple radiological markers of inflammation

To evaluate whether long term treatment with ETI can lead to improvements in lung disease as well as lung function, we performed HRCT scans in 10 modulator naïve patients at baseline and 1 year following therapy.

Those included in this part of the analysis had a spread of lung function at baseline between 45% and 86% predicted at baseline. There was also a spread of lung function response to ETI with 3 showing minimal change (<5% ppFEV1 improvement), 3 showing moderate change (5-15% ppFEV1 improvement) and 4 being “super responders” with >15% improvement in ppFEV1. Their demographics are shown below, with their individual responses to ETI therapy shown in both clinical parameters and systemic inflammatory parameters. Those with minimal change in their ppFEV1 (<5%) are displayed with red dots at 1 year, those with moderate response (5-15%) in ppFEV1 in orange and those with super response (>15%) in green.

Systemic inflammation parameters in the CT cohort are similar to those seen in the larger cohort, with significant reduction in CRP, IL-6, Calprotectin, and monocytes. Neutrophils did not change significantly in this group.

Remarkably CT scans demonstrated improvements in all 10 subjects when qualitatively assessed by 2 independent consultant radiologists. Improvements were seen in all in the qualitative reports in mucus plugging and bronchial wall thickening. Additional improvements were seen in most in bronchiolectasis (small airways disease) and in some with bronchiectasis. A notable finding was a reduction in size significant thoracic lymph nodes in most PWCF, confirmed with a significant reduction in the diameter of their paratracheal lymph nodes (p=0.0002).

Furthermore, PRAGMA scoring by AI analysis using LungQ demonstrated significant improvements across multiple domains including airway wall thickness and total airways disease score. There were no significant changes in inspiratory lung volume as assessed by LungQ despite changes seen in a 13.9% mean improvement in percentage predicted FEV1 and 11.7% predicted improvement in percentage predicted FVC within this cohort. The total airway count did not significantly change, so new airways are not being formed.

### Multiple correlates between systemic inflammation and airway wall thickening are identified following ETI treatment

Utilising the clinical, radiological, inflammatory and basal cell parameters a correlation of the differences seen before and after ETI treatment were assessed. 16 strongly correlated relationships (Pearson R value >0.7 or <-0.7 and p value<0.05) were found and listed below.

CRP, a useful and widely available inflammatory marker was strongly linked with an increase in weight and BMI. It was also strongly correlated with IL-6. Calprotectin, the CF antigen and a biomarker of exacerbation, was strongly correlated with exacerbations.

Surprisingly, age had a significant impact, with younger patients seeing a larger fall in calprotectin, CRP and exacerbations in this group.

In terms of structural lung disease, a reduction in airway wall thickening was strongly linked to paratracheal lymph node size and an improvement in ppFEV1.

**Figure 4A-F:**
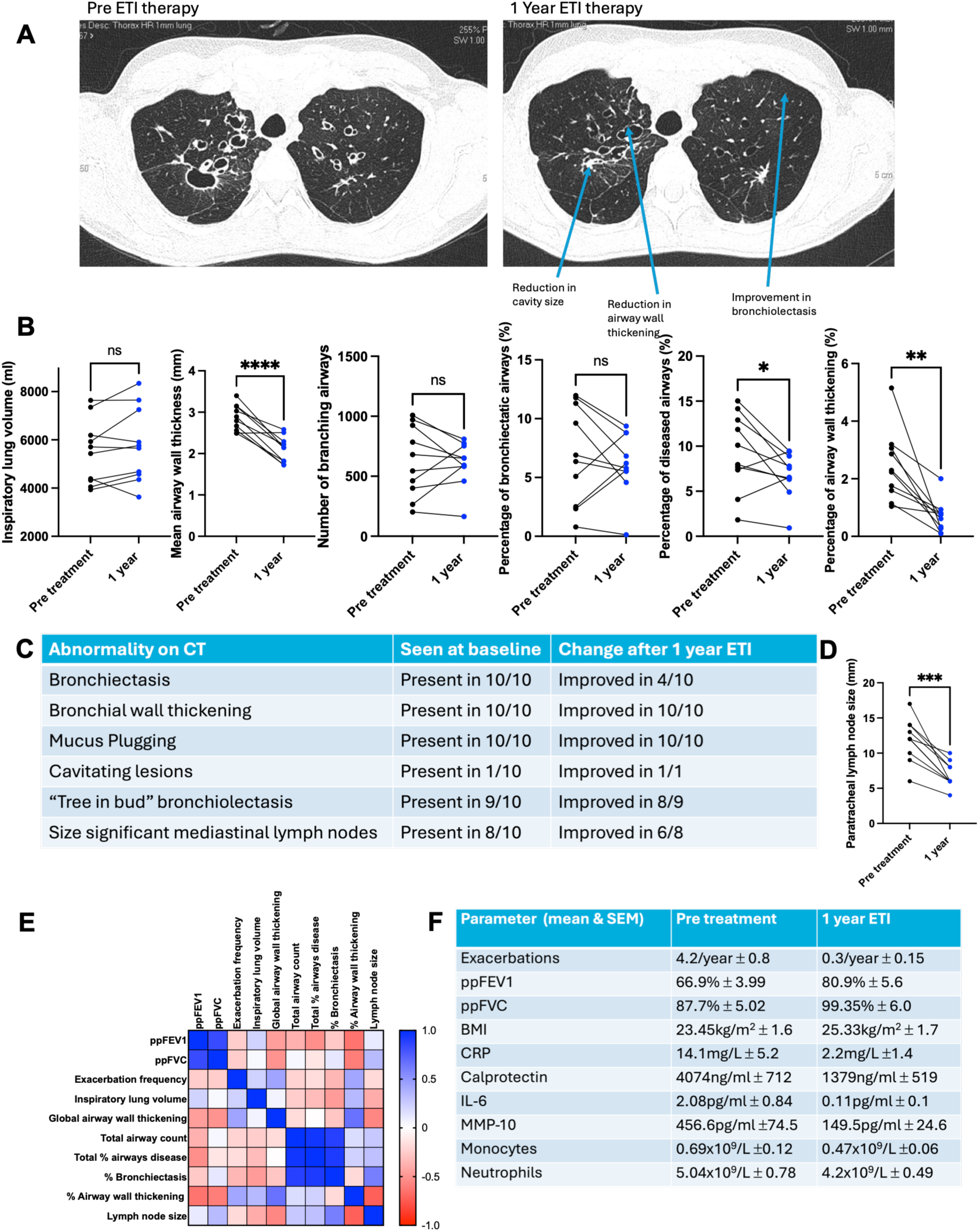
ETI causes significant reduction in radiological markers of pulmonary inflammation, with some changes in structural lung disease. 4A demonstrates a paired cross-sectional high resolution CT thorax image from CF064 prior to commencing ETI and following 1 year of treatment. The arrows show a reduction in the size of an individual cavity, reduction in bronchial wall thickening and reduction in bronchiolectasis. 4B shows the results of the LungQ scans for 10 modulator naïve PWCF pre and post treatment with ETI (n=10). 4C is a table demonstrating the changes reported by independent radiology assessment as a proportion of those scanned (n=10). 4D shows a significant reduction in volume of the diameter of paratracheal lymph nodes across the first year of ETI therapy (n=10). 4E shows a heatmap of correlation of the change in each parameter across the 1 year of treatment, with blue demonstrating a strongly positive correlation and red displaying a strongly negative correlation(n=10). 4F shows a table of the change in clinical and inflammatory parameters across the first year of ETI therapy (n=10) Statistics with paired t tests where *≤0.05, **≤0.01, ***≤0.001, ****≤0.0001 and Pearson R correlates where +1 is blue and -1 is red.

### CF Basal Cells are functionally Different from Non-CF Basal cells and do not form organoids efficiently

**As the resolution of inflammation is essential for tissue repair, we then focussed on the assessment of basal cells from the CF airway the main stem cell population. Firstly, we defined basal cell phenotype in people with CF before CFTR correction, and then following this at multiple time points.** Basal cells were obtained by nasal brushing and selectively expanded from healthy controls and PWCF before and at multiple time points after commencing ETI therapy. To assess how basal cells from PWCF function at baseline we assessed their behaviour in several assays and compared these to healthy basal cells.

Firstly, we measured proliferation by simple cell counting and BRDU assay. Primary CF basal cells replicated at a significantly slower rate than healthy basal cells in culture and exhibited significantly lower BRDU incorporation levels and higher levels of apoptosis albeit below statistical significance.

However, RNA sequencing of both healthy and modulator naïve CF basal cells cultured in control conditions show that these cells have a number of transcriptomic differences. A total of 319 genes were differentially expressed (p.adj < 0.05, absolute log2 fold > 0.5) between the healthy and CF cells at baseline. Gene ontology analysis showed differences in multiple DNA replication pathways, key to their proliferative capacity as airway stem cells.

Functional assessment of basal cells in a wound closure assay demonstrated a non-significant reduction in wound closure at 24 hrs in the CF basal cells (p=0.19). Finally, we assessed organoid formation ex vivo comparing CF to healthy basal cells and showed that CF cells formed significantly smaller organoids by area compared to healthy basal cells (p<0.001).

**Figure 5A-E:**
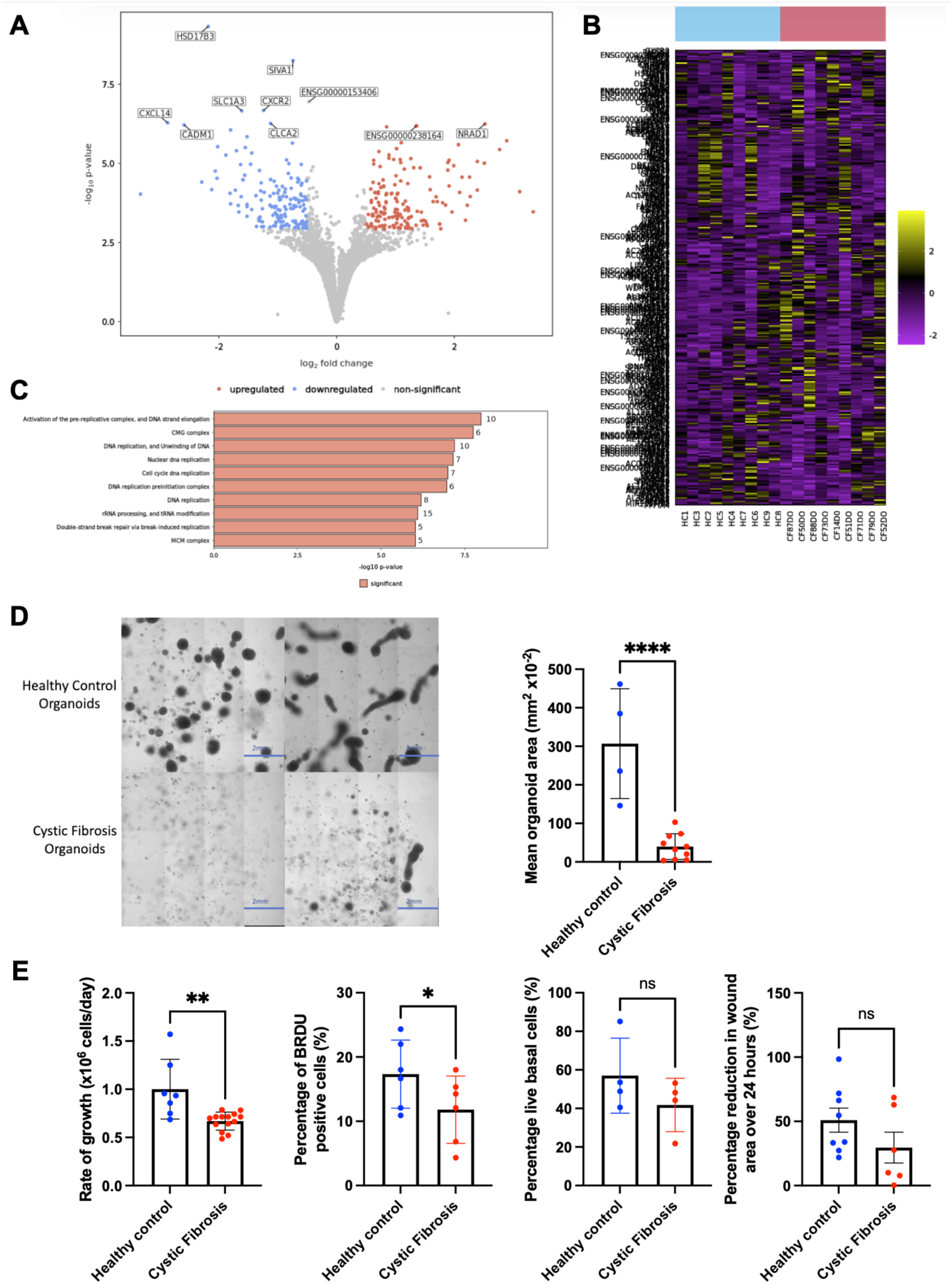
CF basal cells are transcriptomically and functionally different to healthy basal cells. 5A demonstrates the 319 transcriptomic differences between CF and healthy basal cells. Differentially expressed genes (p.adj < 0.05, absolute log2 fold > 0.5) are shown in red (upregulated in CF) and blue (downregulated in CF) and non−significant genes in grey. 5B shows a heatmap of the 100 most significant genes with yellow being upregulated and purple being downregulated. 5C shows the gene ontology top 10 significantly different pathways between CF and healthy basal cells with multiple pathways of DNA replication significantly different. 5D demonstrates 10x magnification stitched light microscopy images of wells containing organoids propagated from healthy and CF basal cells (n=2 healthy and 2 CF) and a significant reduction in the mean organoid area of CF cells compared to healthy controls (n=4healthy and 10 CF). 5E shows a series of experiments of the functional capacity of CF and healthy basal cells. The graphs demonstrate that a significant difference between the growth of healthy and CF basal cells ex vivo when cultured by selective expansion (n= 7 healthy and 13 CF modulator naïve samples), a significant difference in proliferation measured by BRDU between healthy and CF basal cells (n=6 healthy and 6 CF), no significant difference in the rates of apoptosis of healthy or CF cells (n=4 healthy and 4 CF) and CF basal cells are not significantly slower to repair a scratch wound than healthy cells (n= 8 healthy and 6 CF),. Statistics with one way ANOVA with multiple comparisons and t tests where *≤0.05, **≤0.01, ***≤0.001, ****≤0.0001.

### ETI treatment in patients is associated with changes in basal cell gene expression but minimal change in function

PWCF had nasal brushings and basal cells expanded at multiple different time points across the first year of treatment.

CF basal cells improved their rate of growth significantly following 1 month of *in vivo* ETI treatment (p=0.005). However, there is no significant difference at the further *in vivo* timepoints with either ETI, TI or Ivacaftor monotherapy.

To see if this was linked to the *in vivo* changes or the CFTR theraly alone, we utilised CF modulator naïve cells and treated them *in vitro* with CFTR modulators. CFTR modulators *in vitro* did not significantly improve basal cell scratch wound closure on the modulator naïve cells and surprisingly, ETI *in vitro* therapy reduced the size of organoids produced by modulator naïve cells. (p=0.012).

We looked at the transcriptomic signatures of basal cells after 3 months of treatment *in vivo* with ETI and culture *ex vivo* with ETI. This time, CF basal cells had marked changes in basal cell gene expression. 2570 genes were differentially regulated (adjusted p< 0.05, absolute log2 fold >0.5). Gene ontology analysis highlighted. multiple genes associated with cilium assembly were also upregulated. It also included positive regulation of both proliferation and differentiation pathways such as the Notch regulation pathway-important for differentiation of basal cells into their progeny.

**Figure 6A-G:**
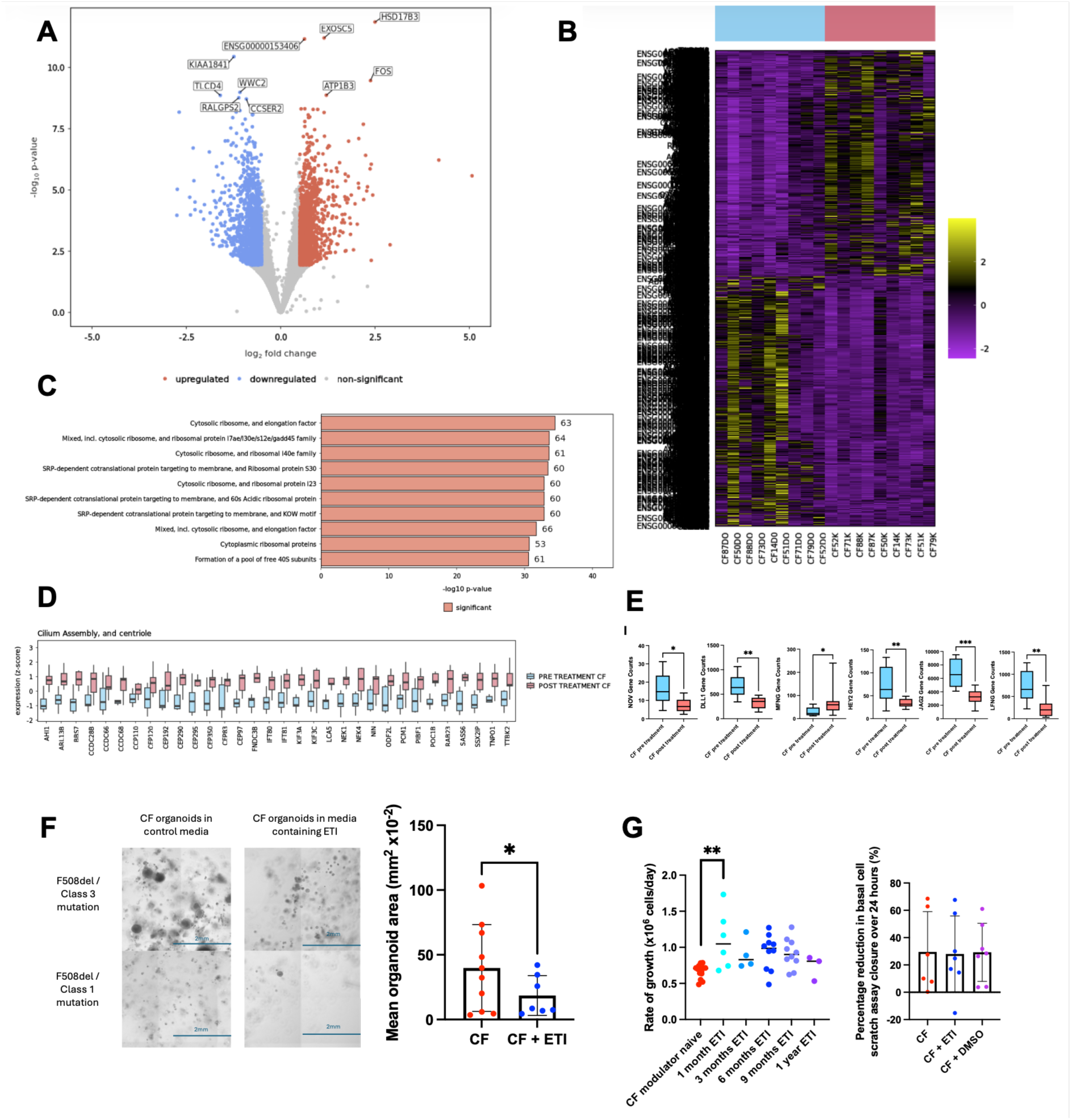
CF basal cells change their transcriptome with ETI therapy but show minimal functional improvement with *ex vivo* ETI treatment. 6A shows the volcano plot of change in CF basal cell transcriptome from RNA sequencing following 3 months of ETI *in vivo* and *ex vivo* ETI supplementation during selective culture of basal cells, 2570 genes were differentially expressed in the paired samples pre and post treatment (n=9) with red dots being upregulated and blue dots downregulated. 6B shows the heatmap of the top 100 significantly different genes between the pre and post treatment groups with yellow being significantly upregulated and purple being significantly downregulated. 6C shows the gene ontology analysis with STRING 11.5 showing the top 10 most significantly different pathways following ETI therapy(n=9). 6D shows the 32 differentially expressed genes related to cilium assembly and centriole found in the basal cells following 3 months of treatment with ETI therapy (n=9). 6E shows differentially expressed Notch signalling pathway genes between pre and post treatment basal cells demonstrating a significant upregulation in Notch (n=9). 6F shows stitched images of 10x bright field magnification of organoids from CF046 and CF075 with and without ETI in vitro (n=2) and a significant reduction in the mean organoid areas for CF modulator naïve basal cells cultured with and without ETI in culture (n=10 without ETI and 7 with ETI). 6G demonstrates the basal cell growth rates *ex vivo* of those who had been treated *in vivo* with CFTR modulators (n=14) and the unchanged wound repair ability of modulator naïve CF basal cells in control media, media supplemented with ETI and that supplemented with DMSO (vehicle control) (n=7). Statistics with one way ANOVA with multiple comparisons and paired t tests where *≤0.05, **≤0.01, ***≤0.001, ****≤0.0001.

## Discussion

CF is caused by genetic mutations in the CFTR gene that lead to lung disease characterised by chronic infection, unremitting inflammation and progressive destructive lung disease^25^. CFTR modulator therapies allow us the opportunity to measure the impact of CFTR correction on both inflammation resolution and tissue repair mechanisms, both of which will be required for long term improvements in respiratory health in people living with CF. The sequential introduction of effective (TI) then highly effective (ETI) offer a unique insight into how the degree of CFTR correction may affects inflammation and consequent tissue repair pathways.

### Impact of TI and ETI on Inflammatory cells

CF is a associated with a hyper-inflammatory immune phenotype with marked abnormalities in neutrophil and macrophage dysfunction^3–511–14,24^, and measureable levels of inflammation both in the lung and systemically. Effective CFTR correction with TI had a limited impact on systemic measurements of inflammation in our cohort with significant improvements only seen in IL-6 after 1 or 3 months of therapy. Interestingly calprotectin, a damage associated molecular pattern (DAMP) which is highly predictive of disease outcomes in CF showed little improvement over the same time course. Likewise, circulating monocytes and neutrophils demonstrated no significant change in phenotype when assessed by multiple cell surface markers with flow cytometry. Taken together these data suggest that effective CFTR correction is not sufficient to supress CF-related inflammation completely although effects on IL-6 may suggest a minor shift in inflammatory phenotype that mirrors the modest effects of TI therapy on lung function.

Based on our preliminary data from TI patients we first looked at the short-term effects of highly effective CFTR correction (ETI) on a panel of serum and cellular measurements of inflammation guided by our work on TI. Despite significant improvements in lung function and other clinical measurements there was only a limited short-term impact of ETI. CRP, IL-6 and calprotectin all significantly decreased over the first three months of treatment, whereas IL-10 and IL-8 levels remained unchanged. Similarly, flow cytometric assessment of circulating neutrophils showed the that significant effects of ETI were limited to increased CD16 staining suggesting increased circulating neutrophil maturity, which may signal lower neutrophil turnover from the bone marrow due to an overall reduction in the inflammatory burden in CF. Significant downward trends in CD47 (don’t eat me signal) and CD35 in addition to the significant rise in CD88 and CD66b would also support the presence of a less activated pool of circulating neutrophils. Similarly circulating monocytes adopted a less classically activated phenotype consistent with an overall shift in the peripheral myeloid population towards a less inflammatory phenotype with highly effective CFTR correction. Taken together with our data demonstrating an overall reduction in circulating myeloid cell numbers, ETI treatment leads to a reduction in both quantity and inflammatory potential of the white blood cells likely to have beneficial effects for people with CF. Our group have previously demonstrated intrinisic features of CFTR deficiency in myeloid cells, independent of the CF inflammatory environment and our present data may represent the effects of CFTR modulators on these intrinsic features as well as the effects of reducing the overall inflammatory burden in the lung as evidenced by others^9,16,25,29^.

### Impact of TI and ETI on inflammatory biomarkers

Next, we assessed the effects of CFTR modulation on inflammatory biomarkers. TI had modest effects on systemic measurements of inflammation with IL-6 being the only systemic marker of inflammation that reduced on effective CFTR correction. In contrast ETI therapy reduced circulating levels of several biomarkers including IL-6, CRP and calprotectin after 3 months of therapy. This is consistent with our cellular data demonstrating a reduction in inflammatory cell numbers but also suggests that inflammation is being impacted at multiple levels. To investigate whether this reduction in inflammatory markers was a global phenomenon we conducted a proteomic analysis of serum inflammation in a group of 10 completely modulator naïve patients before and after 1 year of ETI therapy, to assess the long-term impact of CFTR correction. Interestingly these data demonstrate that out of a panel of 196 inflammatory markers only 3 changed significantly, namely IL-6, IL-20 and MMP10. We then confirmed these findings in our larger cohort for IL-6 and MMP10 (no high sensitivity commercial ELISA available for IL-20). These data suggest that long term therapy is associated with a suppression of IL-6 but also a change in MMP10 a marker of the tissue damage. MMP10 is a member of the matrix metalloproteinase family and is expressed in multiple tissues following injury. It is expressed by macrophages in human lungs of PWCF and can be induced in response to P Aeruginosa infection in mice suggesting it has a beneficial response to acute infection^30^. MMP10 can also clear scar tissues in normal skin and has been shown to promote the polarisation of macrophages towards the repair phenotype, thus limiting the potential of fibrotic responses to injury.^31^ Therefore reductions in MMP10 may signal a reduction in on-going damage in the CF lung. Interestingly IL-6, MMP10 and IL-20 are also components of the senescence associated secretory phenotype (SASP)^32^. SASP represents the response of damaged cells in ageing and chronic inflammatory processes, that although essential for tissue repair, may contribute to inflammaging and tissue dysfunction driving processes such as tumorigenesis. As such a reduction in elements of the SASP phenotype in CF may signal both an improvement in tissue damaging inflammation but also a reduction in the potential risk of SASP which is associated with increased risks of cancer and cardiovascular disease. Furthermore, these individual components of SASP may represent potential therapeutic targets for pWCF unable to take CFTR modulators due to genetics or intolerance.

### The effects of TI and ETI on Calprotectin

The damage-associated molecular pattern (DAMP) protein, calprotectin, has been known to be significantly elevated in the blood, sputum and stool samples of PWCF and can predict exacerbation.^7,8^ It reflects mainly neutrophil activation and is part of the aberrant response to inflammation in multiple organs in CF disease. CFTR modulators statistically significantly reduce this, showing a marked response in serum. We were unable to study calprotectin in sputum following ETI therapy as many of the PWCF stopped producing any spontaneous sputum. However, others who managed to obtain samples showed a reduction in calprotectin in sputum^16^. Our clinical correlation also reveals that this remains strongly linked to exacerbation frequency.

### ETI treatment improves pulmonary inflammation and structural damage in CF

Lung repair is known to be dysfunctional in CF *in vivo*, with development of bronchiectasis through damage to the cartilaginous structures of the airway, basal and goblet cell hyperplasia and a reduction in ciliated cells.^2,21,21,26,33^. We performed serial CT scans on 10 modulator naïve pwCF before and after 1 year of ETI therapy. Remarkably, improvements in radiological appearance were seen in all PWCF in the study across multiple domains including airway wall thickening, mucous plugging and tree and bud bronchiolectasis. This was further quantified in our cohort using automated PRAGMA CT scoring. Our data are consistent with other groups using both the Brody score for calculation, PRAGMA CT scoring.^13,14,34^ Unique to our data set is the observation of a reduction in size significant lymphadenopathy, which may represent an overall reduction in thoracic inflammation with ETI that tracks reductions in other features such as airway wall thickening which itself correlated with improvements in lung function. The improvements in bronchiectasis in some of our cohort suggest the potential for real-life lung tissue repair in CF following modulator treatment, as would be promoted by inflammation resolution. The major stem cell population in the airway is the basal cell and as such we have focussed further experiments on this cell population.

### CF basal cells are genetically and functionally different from non-CF basal cells

When directly sampled from the airway (nasal cavity) CF basal cells have a distinct genetic profile from healthy control samples charactersised by multiple differences in multiple in pathways linked to DNA replication and cell proliferation, suggesting that stem cell capacity may be impaired in CF, although it is unclear whether this is an intrinsic feature of CF basal cells due a decreased CFTR function or indeed secondary to the effects of the inflammatory environment in CF on basal cell function. These transcriptional differences were mirrored by the functional behaviour of CF basal cells which had decreased total proliferative capacity (BRDU incorporation was signifiantly decreased in CF), had slightly poorer wound closure ability (although below the level of statistical significance) and formed organoids less efficiently than non-CF basal cells, suggesting an inability to proliferate and differentiate adequately compared to control tissue.

### Treatment of pwCF with ETI changes CF basal cell transcriptional and functional behavior

ETI treatment for 3 months was associated with significant changes in gene expression across multiple genes and pathways with a particular impact on genes related to ciliagenesis and several pathways associated with cellular proliferation. These data suggest a significant shift in basal cell phenotype towards the production of ciliated cells which are crucial to epithelial function in the airway and have been previously demonstrated as reduced in the CF airway. Interestingly, the functional behaviour of CF basal cells was also improved from pwCF taking ETI with a correction in proliferative capacity. To investigate whether this was a direct effect of CFTR activation by ETI in basal cells we investigate whether the addition of ETI *ex vivo* changed basal cell phenotype or behaviour and demonstrated no impact on proliferation and even a detrimental effect on differentiation into organoids. This may be in part due to the low levels of CFTR recorded in basal cells but also suggest that the environmental effects of CFTR correction in pwCF taking this medication is having the major effect on basal cell phenotype by shifting them towards a reparative population of proliferating basal cells that can perform normal repair functions. Further work is required to evaluate how *in vivo* treatment with ETI impacts basal cell differentiation functions such as the ability to form organoids. Taken together our work suggests that any change in basal cell function in pwCF taking ETI is likely to represent a change in the inflammatory environment in CF impacting cellular function rather than a direct effect of CFTR activation on the basal cell population in isolation. We will investigate this in future studies.

Our data demonstrate that highly effective CFTR modulator treatment significantly reduces specific features of inflammation including the major SASP marker IL-6 and major DAMP calprotectin but has a limited impact on multiple other biomarkers of inflammation. Nevertheless, the clinical impact of this reduction inflammation is evidenced by reduced airway wall thickening on CT in people taking modulators. Furthermore, these reductions in inflammation are associated with transcriptional and functional changes in the major stem cell population from the airway, namely basal cells. And this is likely due to a change in environment rather that the direct effects of CF modulators on basal cells. Further studies are now required to study tissue repair pathways in CF and how these may be augmented to improve lung health in the CF population.

## Conclusions

Although CFTR modulators improve multiple clinical and radiological outcomes, they also have impacts on basal cell function. There are however, limited impacts on systemic inflammation and more work is needed in this area to understand the disease process.

## Data Availability

All data produced in the present study are available upon reasonable request to the authors

